# Hippocampal Volume Predicts Unhealthy Food-Seeking Trajectories in Insulin-Resistant, but Not Insulin-Sensitive, Youth with Obesity and Depression

**DOI:** 10.64898/2026.07.14.26357902

**Authors:** Natalia Khodayari, Jake Branchini, Milong Zhao, Rhesa J. F. Valenzuela, Zachary A. Springs, Maitri Khanna, David Patron, Manpreet K. Singh

**Author notes:** shared first co-authorship (Natalia Khodayari, Jake Branchini).

## Abstract

Insulin resistance, an often-untreated precursor of type 2 diabetes mellitus (T2DM), is implicated in cognitive decline in adults, yet its impact on the developing brain in youth with obesity remains poorly understood. We investigate whether insulin resistance moderates the relation between hippocampal volume and unhealthy food-seeking in overweight and depressed youth ages 9-17 who completed an oral glucose tolerance test and a cognitive task assessing unhealthy food-seeking motivation at baseline, 6-, and 24-months follow-up, and structural MRI at baseline and 6-months follow-up. Insulin sensitivity moderated this relation: smaller baseline hippocampal subfield volumes predicted increased unhealthy food-seeking over 24 months (ps<0.05). Categorical grouping revealed subfield CA2/3 and 4 volumes predicted this relation among insulin-resistant (ps<0.05), but not insulin-sensitive (ps>0.10), youth, suggesting that threshold criteria for insulin resistance are physiologically meaningful. These findings identify a neuro-metabolic risk phenotype that precedes T2DM and may accelerate unhealthy food-seeking severity in youth with obesity.

## Introduction

Rates of obesity in children are rising rapidly. In the United States alone, obesity rates among youth aged 2 to 19 years old have reached a record high of 21.1%. One in five American youth are now obese, with annual medical costs of childhood obesity exceeding $1.3 billion in 2019 (https://www.cdc.gov/obesity/childhood-obesity-facts/childhood-obesity-facts.html). Childhood obesity is associated with various health outcomes, including type 2 diabetes mellitus (T2DM), dyslipidemia, hypertension, asthma, and obstructive sleep apnea.^1^ The prevalence of T2DM in youth is projected to increase substantially over the coming decades,^2^ and childhood obesity increases the risk of depression by 32%.^3^ In the United States, two-thirds of calories consumed by youth come from ultra-processed foods, linking poor diet quality with obesity-related health outcomes.^4^

Insulin resistance in youth with obesity is common but may go undetected as an untreated precursor with neural consequences. Current clinical guidelines from the American Academy of Pediatrics and the American Diabetes Association recommend screening youth aged 10 years or older (or at pubertal onset) with obesity for abnormal glucose metabolism.^5,6^ These screening and treatment frameworks, however, are designed to identify prediabetes and diabetes based on glucose thresholds rather than to characterize insulin resistance itself. Fasting insulin is explicitly not recommended as a reliable measure of insulin resistance,^5^ and OGTT-derived insulin sensitivity indices such as the Matsuda Insulin Sensitivity Index (Matsuda ISI), are not part of routine clinical screening. As a result, youth who are insulin-resistant but have not yet crossed glucose-based diagnostic thresholds for prediabetes may not be identified or targeted for intervention. This gap is particularly important immediately preceding and during adolescence, when key developmental processes support the cognitive regulation of food intake. If obesity and risk for insulin resistance affect this development, they may reinforce unhealthy food-seeking behaviors that contribute to further metabolic dysregulation.

Beyond its peripheral metabolic effects, insulin plays a critical role in the central nervous system, where it facilitates neural circuit formation, synaptic connections, and neuroplasticity from the earliest stages of development.^7^ The hippocampus, a neural region critical for learning, memory, and contextual reward, is rich in insulin receptors and is a key substrate through which dietary patterns and metabolic dysfunction may drive unhealthy eating behavior and obesity risk.^8^ Hippocampal-specific insulin resistance, even in the absence of peripheral metabolic dysfunction, produces structural deficits, impaired spatial learning, and reduced synaptic plasticity^7^ with observed cognitive deficits in both humans and rodent models of T2DM on tasks that rely on the hippocampus.^9^ Adolescents with insulin resistance have been shown to have smaller hippocampal volumes and greater frontal lobe atrophy compared with insulin-sensitive controls.^10^ Administration of intrahippocampal insulin improves performance on hippocampus-dependent memory tasks. In contrast, blocking endogenous insulin signaling within the hippocampus impairs memory processing,^11^ suggesting a causal framework of insulin on hippocampal function.

The hippocampus is also sensitive to signals of hunger and satiety, integrating food-related experiences, environmental cues, and internal physiological signals involved in the regulation of feeding behavior.^8^ In neurologically intact adults, experimentally manipulating the perceived size of a recently consumed meal influences later hunger and expected satiety, demonstrating that hippocampal-dependent meal memory may function as a cognitive brake on eating.^12,13^ Importantly, the relation between the hippocampus and unhealthy food-seeking may be cumulative and structural rather than transient. In a large cross-sectional cohort of 588 children and adolescents, increased BMI was associated with reduced radial thickness in the left anterior hippocampus, a relation driven predominantly by children younger than 14 years, suggesting that childhood may represent a period of particular hippocampal vulnerability to adverse metabolic environments.^14^ Supporting preclinical evidence shows that prolonged high-fat diet consumption during adolescence produces hippocampal neuronal remodeling, microglial overactivation, and synaptic protein loss that worsen with longer exposure duration.^15^

Moreover, the hippocampus integrates inputs with subfield-specific contributions to food-related behavior. Hippocampal *cornu ammonis* subfield CA1 exerts inhibitory control over food-motivated behavior through monosynaptic glutamatergic projections to the medial prefrontal cortex (mPFC); disruption of this ventral CA1-to-mPFC pathway increases food-seeking.^16^ CA1 additionally plays a distinct role in reward valuation, such that inactivation of CA1 impairs incremental value learning during foraging.^17,18^ CA2 neurons support memory consolidation processes that may be relevant to encoding food-reward associations through initiating sharp-wave ripples that propagate to CA3 and CA1.^19^ CA3 parvalbumin-positive inhibitory interneurons in mice additionally play a causal role in food location-learning: these CA3 neurons reduce their firing around novel food locations, enabling the CA3 network to form memories which link spatial locations to food rewards. Disruption of this memory process results in mice failing to learn where to find food.^20^ In children, decreased CA4 (i.e., hilus of the dentate gyrus) grey matter volume have been associated with loss-of-control eating at an uncorrected threshold, a pattern consistent with binge eating disorder risk.^21^

Together, hippocampal circuitry is highly metabolic and directly implicated in the development of food-seeking behaviors. These findings suggest a reinforcing loop in which insulin resistance driven by chronic unhealthy food intake contributes to progressive hippocampal structural compromise, which may in turn impair the hippocampal-dependent memory and interoceptive processes that normally regulate food intake, making later intake more difficult to control.^13^ Thus, disruption of insulin dynamics may affect hippocampal-dependent food regulation mechanisms and further reinforce unhealthy food-seeking.^8,22^ Our group has previously reported that depressed and overweight or obese youth aged 9 to 17 with greater insulin resistance have smaller hippocampal volumes, reduced anterior cingulate cortex (ACC) volumes, and increased food-seeking behavior.^23^ Insulin resistance in this cohort was also associated with smaller whole brain volumes independent of BMI, depression severity, and IQ.^24^ Yet it remains unclear (1) whether structural differences in the hippocampus in youth may predict development of unhealthy food-seeking behaviors over time, and (2) whether individual differences in insulin sensitivity modulate the relation between hippocampal structure and food-seeking trajectories in youth.

These can be evaluated in the same youth who, in addition, to having baseline structural MRI, also completed an adaptation of the Relative Reinforcing Value (RRV) food-seeking cognitive task^25^ with 6- and 24-month longitudinal follow-up to objectively assess food-seeking behavior.

We hypothesized that hippocampal volumes would predict changes in unhealthy food-seeking trajectories, such that smaller hippocampal subfield CA1-4 volumes at baseline would predict greater unhealthy food-seeking over time in youth. We further hypothesized that insulin sensitivity would moderate the relation between hippocampal volume and food-seeking trajectories, such that this relation would be stronger among youth less sensitive to insulin (i.e., more insulin resistant), reflecting impaired food-related inhibitory and mnemonic processes that regulate food intake. Insulin sensitivity is operationalized using Matsuda ISI both continuously and grouped categorically via median split aligned with proposed clinical thresholds of insulin resistance using Matsuda ISI.

## Results

See Table 1 for demographics. Test-retest analyses (Fig. 1) demonstrate robust test-retest reliability of unhealthy food-seeking across timepoints with Bayesian evidence supporting this relation.

**Fig. 1.**
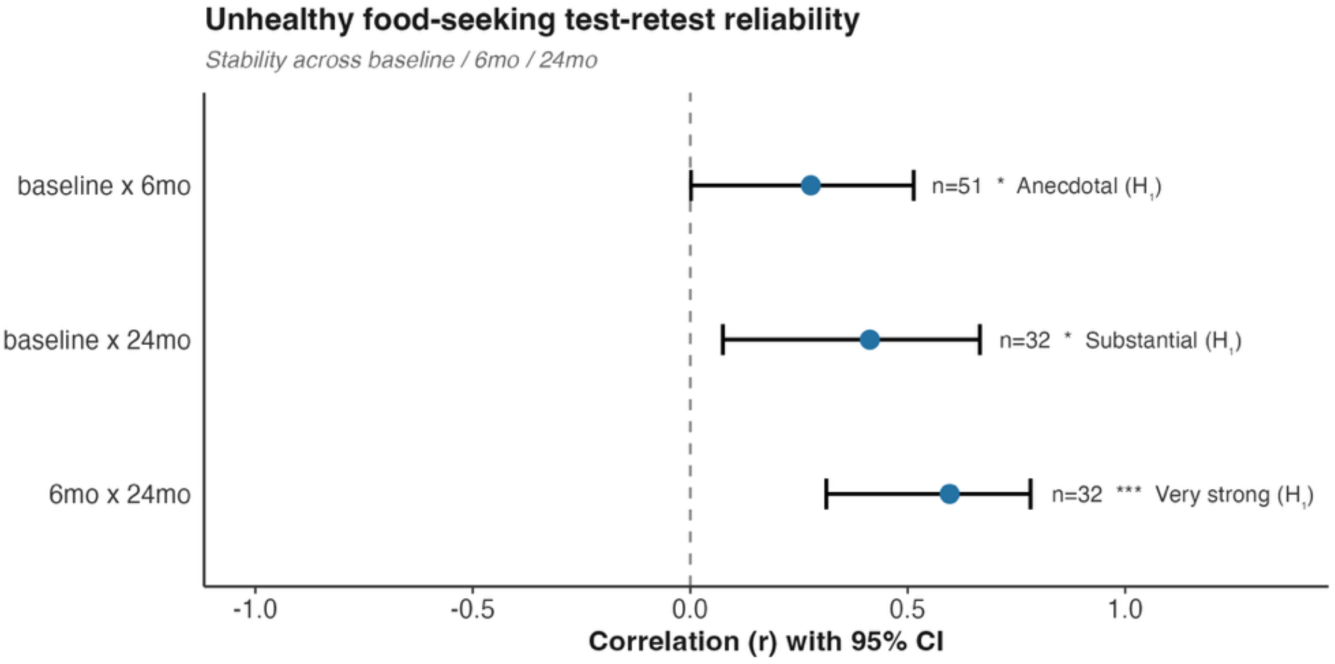
Reliability and Stability of Unhealthy Eating. Pearson test-retest correlation comparisons of unhealthy food seeking between: baseline vs 6-months follow-up (top row; p=0.049), baseline vs 24-month follow-up (middle row; p= 0.019), and 6- vs 24-month follow-up (bottom row; p=0.0003). Blue point reflects correlation coefficient (r). Horizontal bars reflect 95% confidence interval. Sample size (n), significance (*p<0.05, **p<0.01, ***p<0.001), and Bayes Factor (BF_10_) are reported to the right of each comparison. Vertical dashed line reflects correlation coefficient of 0 for reference.

**Table 1.**
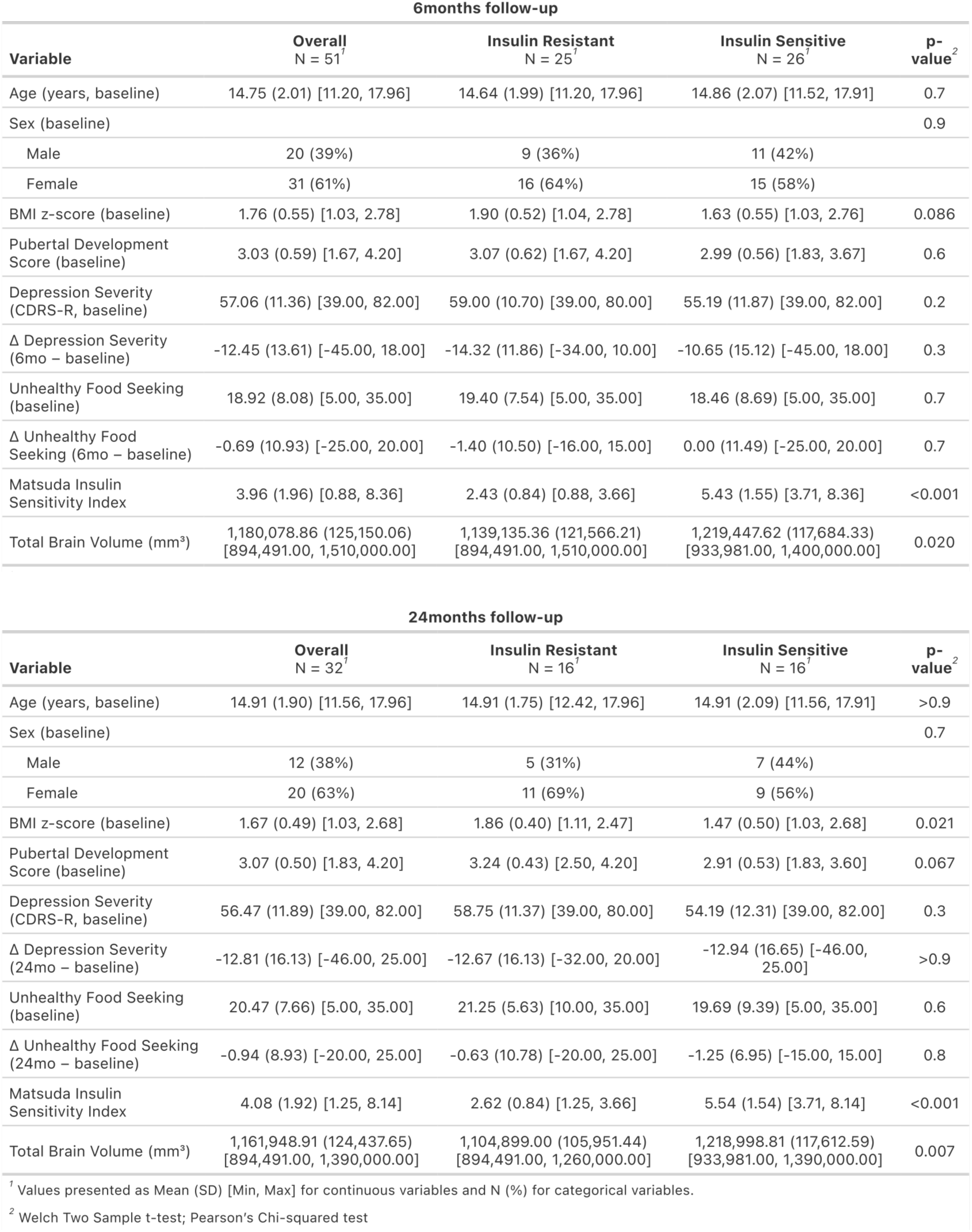
Study Variables.

| 6months follow-up |  |  |  |  |
| --- | --- | --- | --- | --- |
| Variable | Overall<br>N = 51<sup>1</sup> | Insulin Resistant<br>N = 25<sup>1</sup> | Insulin Sensitive<br>N = 26<sup>1</sup> | p-value<sup>2</sup> |
| Age (years, baseline) | 14.75 (2.01) [11.20, 17.96] | 14.64 (1.99) [11.20, 17.96] | 14.86 (2.07) [11.52, 17.91] | 0.7 |
| Sex (baseline) |  |  |  | 0.9 |
| Male | 20 (39%) | 9 (36%) | 11 (42%) |  |
| Female | 31 (61%) | 16 (64%) | 15 (58%) |  |
| BMI z-score (baseline) | 1.76 (0.55) [1.03, 2.78] | 1.90 (0.52) [1.04, 2.78] | 1.63 (0.55) [1.03, 2.76] | 0.086 |
| Pubertal Development Score (baseline) | 3.03 (0.59) [1.67, 4.20] | 3.07 (0.62) [1.67, 4.20] | 2.99 (0.56) [1.83, 3.67] | 0.6 |
| Depression Severity (CDRS-R, baseline) | 57.06 (11.36) [39.00, 82.00] | 59.00 (10.70) [39.00, 80.00] | 55.19 (11.87) [39.00, 82.00] | 0.2 |
| Δ Depression Severity (6mo – baseline) | -12.45 (13.61) [-45.00, 18.00] | -14.32 (11.86) [-34.00, 10.00] | -10.65 (15.12) [-45.00, 18.00] | 0.3 |
| Unhealthy Food Seeking (baseline) | 18.92 (8.08) [5.00, 35.00] | 19.40 (7.54) [5.00, 35.00] | 18.46 (8.69) [5.00, 35.00] | 0.7 |
| Δ Unhealthy Food Seeking (6mo – baseline) | -0.69 (10.93) [-25.00, 20.00] | -1.40 (10.50) [-16.00, 15.00] | 0.00 (11.49) [-25.00, 20.00] | 0.7 |
| Matsuda Insulin Sensitivity Index | 3.96 (1.96) [0.88, 8.36] | 2.43 (0.84) [0.88, 3.66] | 5.43 (1.55) [3.71, 8.36] | <0.001 |
| Total Brain Volume (mm<sup>3</sup>) | 1,180,078.86 (125,150.06) [894,491.00, 1,510,000.00] | 1,139,135.36 (121,566.21) [894,491.00, 1,510,000.00] | 1,219,447.62 (117,684.33) [933,981.00, 1,400,000.00] | 0.020 |
| 24months follow-up |  |  |  |  |
| Variable | Overall<br>N = 32<sup>1</sup> | Insulin Resistant<br>N = 16<sup>1</sup> | Insulin Sensitive<br>N = 16<sup>1</sup> | p-value<sup>2</sup> |
| Age (years, baseline) | 14.91 (1.90) [11.56, 17.96] | 14.91 (1.75) [12.42, 17.96] | 14.91 (2.09) [11.56, 17.91] | >0.9 |
| Sex (baseline) |  |  |  | 0.7 |
| Male | 12 (38%) | 5 (31%) | 7 (44%) |  |
| Female | 20 (63%) | 11 (69%) | 9 (56%) |  |
| BMI z-score (baseline) | 1.67 (0.49) [1.03, 2.68] | 1.86 (0.40) [1.11, 2.47] | 1.47 (0.50) [1.03, 2.68] | 0.021 |
| Pubertal Development Score (baseline) | 3.07 (0.50) [1.83, 4.20] | 3.24 (0.43) [2.50, 4.20] | 2.91 (0.53) [1.83, 3.60] | 0.067 |
| Depression Severity (CDRS-R, baseline) | 56.47 (11.89) [39.00, 82.00] | 58.75 (11.37) [39.00, 80.00] | 54.19 (12.31) [39.00, 82.00] | 0.3 |
| Δ Depression Severity (24mo – baseline) | -12.81 (16.13) [-46.00, 25.00] | -12.67 (16.13) [-32.00, 20.00] | -12.94 (16.65) [-46.00, 25.00] | >0.9 |
| Unhealthy Food Seeking (baseline) | 20.47 (7.66) [5.00, 35.00] | 21.25 (5.63) [10.00, 35.00] | 19.69 (9.39) [5.00, 35.00] | 0.6 |
| Δ Unhealthy Food Seeking (24mo – baseline) | -0.94 (8.93) [-20.00, 25.00] | -0.63 (10.78) [-20.00, 25.00] | -1.25 (6.95) [-15.00, 15.00] | 0.8 |
| Matsuda Insulin Sensitivity Index | 4.08 (1.92) [1.25, 8.14] | 2.62 (0.84) [1.25, 3.66] | 5.54 (1.54) [3.71, 8.14] | <0.001 |
| Total Brain Volume (mm<sup>3</sup>) | 1,161,948.91 (124,437.65) [894,491.00, 1,390,000.00] | 1,104,899.00 (105,951.44) [894,491.00, 1,260,000.00] | 1,218,998.81 (117,612.59) [933,981.00, 1,390,000.00] | 0.007 |
| <sup>1</sup> Values presented as Mean (SD) [Min, Max] for continuous variables and N (%) for categorical variables. |  |  |  |  |
| <sup>2</sup> Welch Two Sample t-test; Pearson's Chi-squared test |  |  |  |  |

Using a median split to generate insulin sensitivity groups (median split: 3.71; below median=Insulin Resistant; above median=Insulin Sensitive) (Fig. 2), we found an average Matsuda ISI of 2.43 at baseline for the Insulin Resistant (IR) group and 5.43 for the Insulin Sensitive (IS) group, confirming alignment with measures observed in clinical populations with insulin resistance (see Methods for more details).

**Fig. 2.**
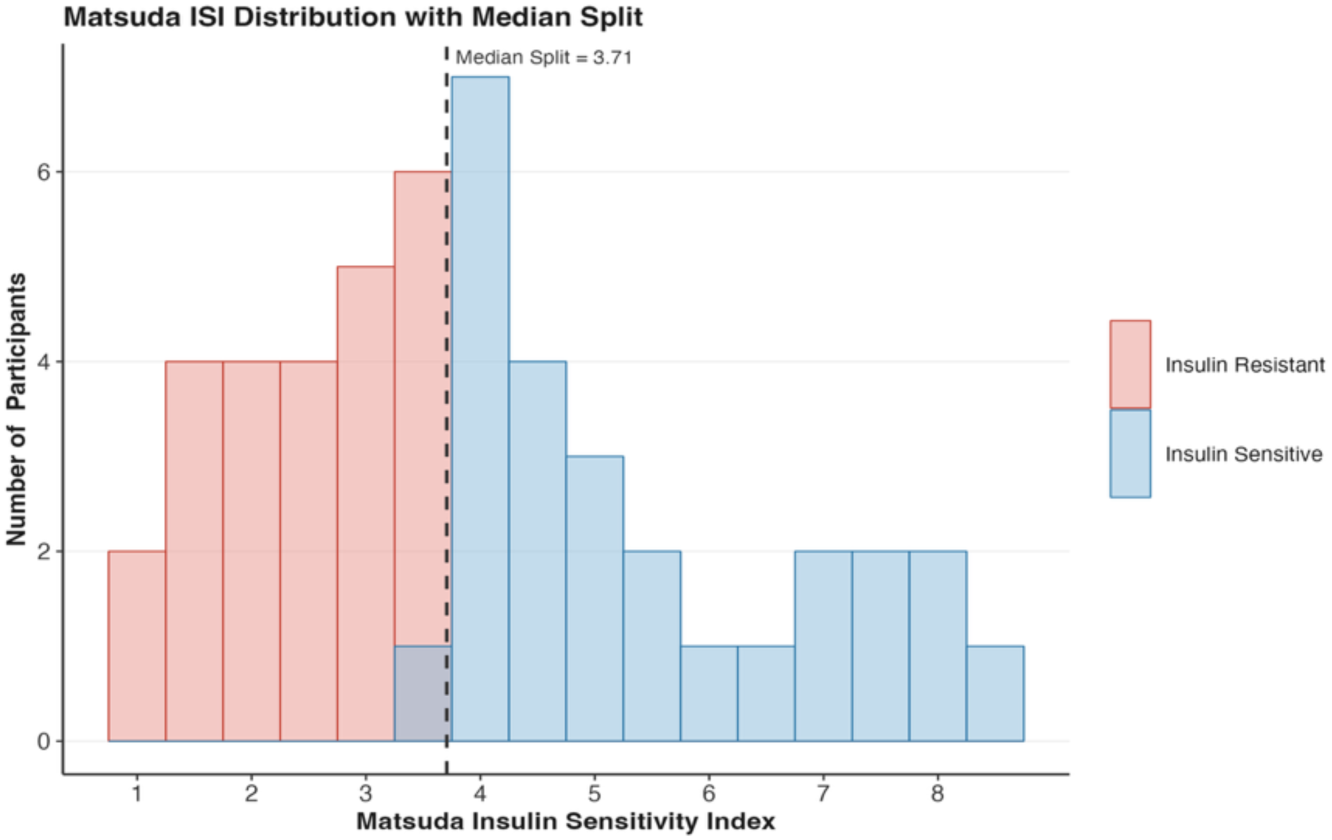
Matsuda ISI Distribution and Median Split. Histogram of Matsuda ISI distribution across participants. Median split (Matsuda ISI=3.71) is indicated by a vertical black dashed line with groups indicated by color (Insulin Resistant group=red bars, Insulin Sensitive group=blue bars). Range of Matsuda ISI across participants and groups is reported in Table 1.

**Fig. 3.**
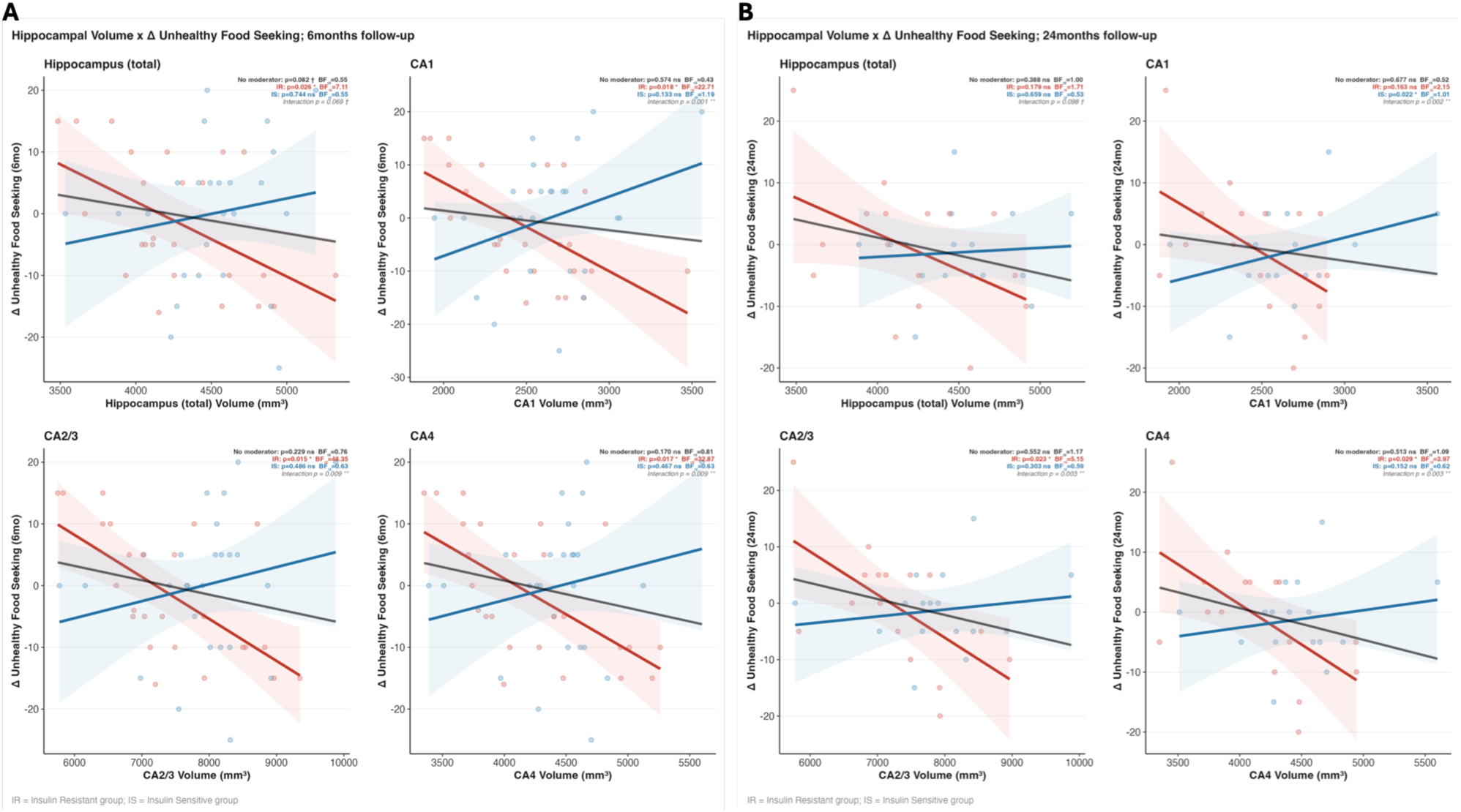
Correlation comparison plots. Moderation of Insulin Sensitivity on the relation between hippocampal volume at baseline and change in unhealthy food-seeking from 6-months – baseline (A) and 24-months – baseline (B). Plots depict unadjusted simple slopes with displayed statistical values reflecting addition of covariates. All statistics reported reflect methods described in Methods. Red line and text: Insulin Resistant group; Blue line and text: Insulin Sensitive group. For each comparison, the group effect p, Bayes Factor (BF_10_), and interaction p are presented in the top right side of the comparison plot. We present four comparisons per timepoint using the following regions of interest (x-axis): average hippocampal volume (top left), and hippocampal subfields CA1 (top right), CA2/3 (bottom left), and CA4 (bottom right). Black line: model fit without insulin sensitivity moderation (all p > 0.05).

**Fig. 4.**
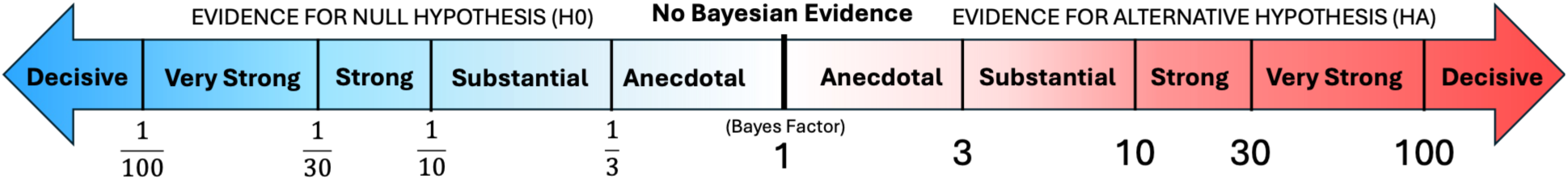
Bayes Factor scale. Bayes Factor greater than 1 provides evidence in favor of the alternative hypothesis (HA). In this study evidence in favor of the HA means evidence in favor of a relation between hippocampal volume and change in unhealthy food-seeking. Bayes Factor less than 1 provides evidence in favor of the null hypothesis (H0) . In this study evidence in favor of the HA means evidence in favor of no relation between hippocampal volume and change in unhealthy food-seeking.

### Continuous Insulin Sensitivity Moderation

Without insulin sensitivity moderation, we found no significant relation between average bilateral hippocampal volume at baseline and change in unhealthy food-seeking at 6- and 24-month follow-up (all p > 0.05). Individual differences of insulin sensitivity, however, significantly moderated this relation at 6- month follow-up (p < 0.05) and non-significant, but marginal, at 24-month follow-up (p=0.054), such that as insulin sensitivity decreases (i.e., insulin resistance increases) the relation between hippocampal volume and change in unhealthy food-seeking becomes more negative. Bayesian analyses provided anecdotal support for this relation at 6- and 24-month follow-up (6-month: BF_10_=2.995; 24-month: BF_10_=2.36).

Across hippocampal subfields CA1, CA2/3, and CA4, we found no significant relation between subfield volumes at baseline and change in unhealthy food-seeking at 6- and 24-month follow-up (all p > 0.05). Insulin sensitivity, however, significantly moderated the relation between subfield volume and unhealthy food-seeking at both 6- and 24-month follow-ups (all p < 0.05) and remained predictive following Benjamini-Hochberg FDR correction for multiple comparisons at 6- and 24-month follow-up (all p_BH_ < 0.05). Substantial to very strong Bayesian evidence was in support of this moderated relation at 6- and 24-month follow-up, respectively (6-month: all BF_10_ > 24; 24-month: all BF_10_ > 6).

### Categorical Insulin Sensitivity Moderation

Overall bilateral hippocampal volume at baseline significantly predicted changes in unhealthy food-seeking (RRV task) from baseline to 6-months among participants in the IR group (p=0.026), such that smaller hippocampal volumes predicted greater increases in food-seeking. This relation was not observed at 24-months follow-up (p=0.18) or among participants in the IS group (6-month: p=0.74; 24- month: p=0.66). The interaction between insulin sensitivity group and hippocampal volume was non-significant at 6- and 24-months follow-up (6-months: marginal, p=0.069; 24-months: p=0.098). At 6- months, we found substantial Bayesian evidence in support of the relation between hippocampal volume and unhealthy food-seeking (BF_10_=7.11) in the IR group and anecdotal Bayesian evidence for no association between hippocampal volume and change in unhealthy food-seeking in the IS group (BF_10_=0.55). At 24-months we found anecdotal Bayesian evidence for an association in the IR group (BF_10_=1.71) and for no association in the IS group (BF_10_=0.53).

Baseline volumes for hippocampal subfields CA2/3, and CA4 were significantly predictive of change in unhealthy food-seeking in the IR group (all ps < 0.05) and remained predictive following Benjamini-Hochberg FDR correction for multiple comparisons (all p_BH_ < 0.05) across both 6- and 24- months follow-up with significant interaction (all p_BH_ < 0.05), such that smaller hippocampal subfield volumes at baseline predicted increases in unhealthy food over 6- and 24-months for IR but not IS groups (all p > 0.05). We found substantial-to-strong Bayesian evidence in support of this relation (6-month: all BF_10_ > 22; 24-month: all BF_10_ > 3). Bayesian evidence in the IS group anecdotally supported this relation to CA1 (BF_10_=1.2) and anecdotally supported no relation to CA2/3 and CA4 (BF_10_ < 0.63).

Subfield CA1 was significantly predictive of this relation at 6-months in the IR (p=0.018; p_BH_=0.018), not IS (p=0.133), group with significant interaction (p=0.001; p_BH_=0.004). CA1 volume at 24-months, however, was associated with change in food-seeking only in the IS (p=0.022), not IR (p=0.163) group with significant interaction (p=0.002). This relation in the IS group was non-significant, but marginal, following FDR correction (p_BH_=0.06) with a significant interaction between groups (p_BH_=0.004). We additionally found anecdotal Bayesian evidence in support of this relation (IS group: BF_10_=1.01; IR group: BF_10_=2.15).

We examined post hoc whether the relation between baseline hippocampal volume and change in unhealthy food-seeking could be instead explained by change in depression severity, as our cohort was originally recruited for comorbid obesity and depression. We found that our results could not be explained by individual differences in depression severity (see Supplemental Materials).

### Peripheral Insulin Exploration

As the Matsuda ISI captures whole body (central and peripheral) insulin sensitivity, we explored whether our results could be explained by the following peripheral insulin factors: HOMA-IR (hepatic insulin resistance), insulin AUC (total insulin secretion), fasting insulin (basal insulin secretion during fasting), insulinogenic index (early stage β-cell insulin secretion). We found that across both 6- and 24- month trajectories, our original findings for the IR group remained significant after including each respective peripheral insulin factor into the primary model, i.e. equation (3), separately (all ps < 0.05) and when all peripheral factors were added to the model (all ps<0.05) (see Table 2). The relation between hippocampal subregion CA1 and change in unhealthy food-seeking at 24-months follow-up became significant after adding insulinogenic Index or Insulin AUC to the model, such that smaller hippocampal CA1 volumes predicted increased unhealthy food-seeking at 24-months follow-up for participants in the IR group (ps<0.05). All relations between hippocampal volume and change in unhealthy food-seeking in the IS group remained non-significant, and the CA1 subfield at 24-months became non-significant (all ps > 0.05).

**Table 2.** Statistics for exploratory covariate analyses. Each covariate column reports the coefficient and p-value for hippocampal volume (β, p) within the given insulin-sensitivity group (top: Insulin Resistant; bottom: Insulin Sensitive), from equation (3) after separately adding the given peripheral covariate. The ‘Combined’ column reflects the same coefficient after adding all peripheral covariates (i.e., HOMA-IR, Insulin AUC, Fasting Insulin, & Insulinogenic Index) simultaneously. **†** p < .10, **\*** p < 00.05, **\*\*** p < 00.01, **\*\*\*** p < 00.001.

| Peripheral Metabolic Covariate Exploration |  |  |  |  |  |  |  |
| --- | --- | --- | --- | --- | --- | --- | --- |
| ROI | Timepoint | Primary model | HOMA-IR | Insulin AUC | Fasting Insulin | Insulinogenic Index | Combined |
| **Insulin Resistant** |  |  |  |  |  |  |  |
| Hippocampus (total) | 6 months | $\beta = -0.017, p = .026 *$ | $\beta = -0.017, p = .027 *$ | $\beta = -0.021, p = .042 *$ | $\beta = -0.017, p = .027 *$ | $\beta = -0.021, p = .010 *$ | $\beta = -0.022, p = .030 *$ |
| CA1 | 6 months | $\beta = -0.018, p = .018 *$ | $\beta = -0.018, p = .017 *$ | $\beta = -0.021, p = .028 *$ | $\beta = -0.018, p = .017 *$ | $\beta = -0.018, p = .026 *$ | $\beta = -0.019, p = .042 *$ |
| CA2/3 | 6 months | $\beta = -0.007, p = .015 *$ | $\beta = -0.007, p = .013 *$ | $\beta = -0.007, p = .033 *$ | $\beta = -0.007, p = .012 *$ | $\beta = -0.007, p = .015 *$ | $\beta = -0.007, p = .035 *$ |
| CA4 | 6 months | $\beta = -0.012, p = .017 *$ | $\beta = -0.012, p = .017 *$ | $\beta = -0.013, p = .034 *$ | $\beta = -0.012, p = .016 *$ | $\beta = -0.012, p = .016 *$ | $\beta = -0.013, p = .029 *$ |
| Hippocampus (total) | 24 months | $\beta = -0.011, p = .179 \text{ ns}$ | $\beta = -0.011, p = .244 \text{ ns}$ | $\beta = -0.023, p = .085 \dagger$ | $\beta = -0.010, p = .249 \text{ ns}$ | $\beta = -0.018, p = .064 \dagger$ | $\beta = -0.025, p = .140 \text{ ns}$ |
| CA1 | 24 months | $\beta = -0.011, p = .163 \text{ ns}$ | $\beta = -0.011, p = .206 \text{ ns}$ | $\beta = -0.026, p = .035 *$ | $\beta = -0.011, p = .199 \text{ ns}$ | $\beta = -0.020, p = .049 *$ | $\beta = -0.024, p = .132 \text{ ns}$ |
| CA2/3 | 24 months | $\beta = -0.008, p = .023 *$ | $\beta = -0.009, p = .026 *$ | $\beta = -0.013, p = .010 **$ | $\beta = -0.008, p = .031 *$ | $\beta = -0.009, p = .018 *$ | $\beta = -0.016, p = .032 *$ |
| CA4 | 24 months | $\beta = -0.012, p = .029 *$ | $\beta = -0.013, p = .034 *$ | $\beta = -0.020, p = .016 *$ | $\beta = -0.012, p = .039 *$ | $\beta = -0.015, p = .023 *$ | $\beta = -0.028, p = .012 *$ |
| **Insulin Sensitive** |  |  |  |  |  |  |  |
| Hippocampus (total) | 6 months | $\beta = -0.003, p = .744 \text{ ns}$ | $\beta = -0.004, p = .682 \text{ ns}$ | $\beta = -0.006, p = .542 \text{ ns}$ | $\beta = -0.004, p = .668 \text{ ns}$ | $\beta = -0.005, p = .558 \text{ ns}$ | $\beta = -0.006, p = .518 \text{ ns}$ |
| CA1 | 6 months | $\beta = 0.011, p = .133 \text{ ns}$ | $\beta = 0.011, p = .155 \text{ ns}$ | $\beta = 0.008, p = .351 \text{ ns}$ | $\beta = 0.011, p = .155 \text{ ns}$ | $\beta = 0.009, p = .265 \text{ ns}$ | $\beta = 0.008, p = .358 \text{ ns}$ |
| CA2/3 | 6 months | $\beta = 0.002, p = .486 \text{ ns}$ | $\beta = 0.002, p = .590 \text{ ns}$ | $\beta = 0.001, p = .792 \text{ ns}$ | $\beta = 0.002, p = .603 \text{ ns}$ | $\beta = 0.001, p = .710 \text{ ns}$ | $\beta = 0.001, p = .865 \text{ ns}$ |
| CA4 | 6 months | $\beta = 0.004, p = .467 \text{ ns}$ | $\beta = 0.004, p = .543 \text{ ns}$ | $\beta = 0.002, p = .794 \text{ ns}$ | $\beta = 0.004, p = .561 \text{ ns}$ | $\beta = 0.003, p = .669 \text{ ns}$ | $\beta = 0.001, p = .841 \text{ ns}$ |
| Hippocampus (total) | 24 months | $\beta = 0.004, p = .659 \text{ ns}$ | $\beta = 0.004, p = .667 \text{ ns}$ | $\beta = 0.003, p = .805 \text{ ns}$ | $\beta = 0.004, p = .667 \text{ ns}$ | $\beta = 0.004, p = .688 \text{ ns}$ | $\beta = 0.002, p = .872 \text{ ns}$ |
| CA1 | 24 months | $\beta = 0.017, p = .022 *$ | $\beta = 0.017, p = .025 *$ | $\beta = 0.017, p = .034 *$ | $\beta = 0.017, p = .026 *$ | $\beta = 0.018, p = .018 *$ | $\beta = 0.015, p = .074 \dagger$ |
| CA2/3 | 24 months | $\beta = 0.003, p = .303 \text{ ns}$ | $\beta = 0.003, p = .300 \text{ ns}$ | $\beta = 0.003, p = .423 \text{ ns}$ | $\beta = 0.003, p = .309 \text{ ns}$ | $\beta = 0.003, p = .290 \text{ ns}$ | $\beta = 0.002, p = .489 \text{ ns}$ |
| CA4 | 24 months | $\beta = 0.008, p = .152 \text{ ns}$ | $\beta = 0.009, p = .151 \text{ ns}$ | $\beta = 0.008, p = .185 \text{ ns}$ | $\beta = 0.008, p = .162 \text{ ns}$ | $\beta = 0.009, p = .130 \text{ ns}$ | $\beta = 0.007, p = .212 \text{ ns}$ |

## Discussion

These findings demonstrate that insulin sensitivity moderates developmental trajectories between baseline hippocampal volume and unhealthy food-seeking in youth with obesity and moderate-to-severe depression. Smaller average bilateral hippocampal volumes and subfield CA1, CA2/3, and CA4 volumes at baseline predicted increased unhealthy food-seeking at 6-months follow-up, but only among youth meeting criteria for insulin resistance. Subfield analyses localized this effect to CA2/3 and CA4 which persisted at 24 months, suggesting that these relations are driven by subfield-specific processes rather than global changes in hippocampal volume.

This work builds on prior studies of insulin resistance and food-seeking behavior which have primarily identified cross-sectional associations,^23^ dose-response relations between glucose metabolism and hippocampal subfield volumes,^26^ and associations between hippocampal volume to insulin sensitivity scores in adults with T2DM.^27^ This is the first longitudinal study, however, to link insulin sensitivity and hippocampal subfields to predict unhealthy food-seeking trajectories in adolescents with obesity and depression. Consistent with our findings, the Maastricht Study (n=4,724) found that T2DM was associated with smaller volumes specifically in CA2/3, and CA4, with continuous hyperglycemia measures showing the strongest associations with CA3 volumes, further supporting the metabolic vulnerability of these subfields.^26^ Given that current screening frameworks focus on glucose-based thresholds for prediabetes and diabetes rather than on insulin resistance directly, our results identify a population of youth whose untreated insulin resistance may drive neural changes with maladaptive cognitive and behavioral outcomes and result in serious downstream metabolic consequences.

Several converging lines of evidence suggest distinct mechanisms across hippocampal subfields which may explain why smaller hippocampal volumes predict greater food-seeking behavior in insulin resistant youth as we have found. First, insulin plays a critical role in regulating hippocampal synaptic plasticity, neurogenesis, and memory-related cognitive functions such as spatial learning and contextual memory.^28^ The hippocampus is rich in insulin receptors and has a particularly slow insulin receptor turnover rate compared to other insulin receptor-dense regions.^29^ Insulin resistance may therefore attenuate hippocampal neural plasticity and neurogenesis by reducing insulin binding in a region where receptor replenishment is already limited, contributing to reduced hippocampal volume and diminished capacity for the inhibitory appetitive and food-related mnemonic processes critical for healthy food-seeking behaviors. These metabolic and neuronal changes may generate a positive feedback loop, such that hippocampal plasticity and volume reductions driven by insulin resistance may result in worsening unhealthy food-seeking, which may further worsen insulin resistance and therefore continue to exacerbate hippocampal plasticity and volume reductions. Second, the hippocampus exerts inhibitory control over food-motivated behavior through projections to the medial prefrontal cortex (mPFC). Hsu et al.^16^ demonstrated that a monosynaptic glutamatergic ventral CA1-to-mPFC pathway inhibits food-motivated behaviors through GLP-1 receptor signaling, such that disruption of this pathway increases operant responding for palatable food. This CA1-specific mechanism may contribute to the transient CA1 association observed at 6 months in the Insulin Resistant group, while the persistent CA2/3 and CA4 effects may instead reflect distinct food-location memory and pattern separation processes. Third, the hippocampus integrates episodic meal-related memories with interoceptive signals of hunger and satiety to regulate subsequent food intake.^8^ Given that CA3 neurons encode food-location memories through parvalbumin-positive interneuron-mediated learning,^20^ and that CA2-initiated sharp-wave ripples support the consolidation of these food-reward associations,^19^ impaired hippocampal function in these subfields may subsequently reduce one’s ability to use prior meal memories to suppress appetite, leading to increased food-seeking. The persistence of CA2/3 and CA4 effects on food seeking at both 6- and 24- months is consistent with disruption of these mnemonic processes.

The localization of durable unhealthy food-seeking trajectories to subfields CA2/3 and CA4 is supported by prior research examining food-related processing. Memory tasks guided by food reward are shown to enhance hippocampal high-frequency neural oscillations (i.e., sharp wave-ripples) found in CA2 and CA3 supporting memory consolidation and foraging decisions.^30^ Distinct neural populations in the dorsal hippocampus show increased activity following the intake of fats or sugars, and contribute to encoding memory for the location and intake of food.^31^ Inhibition of dopamine-2 receptor neurons in the dorsal CA3 region of the hippocampus increased food intake.^32^ Decreased grey matter volumes of CA4 have been associated with loss-of-control eating in children at an uncorrected threshold.^21^ The Maastricht Study further demonstrated that continuous hyperglycemia measures were most strongly associated with CA3 and dentate gyrus volumes in adults,^26^ consistent with the metabolic sensitivity of these subfields observed in our adolescent cohort. CA1 was significantly predictive of food-seeking trajectories in the IR group at 6 months but did not persist at 24 months, which was a distinct temporal pattern from CA2/3 and CA4. This transient association is consistent with the established role of CA1 in food-reward valuation and inhibitory output to the mPFC,^16,17^ which may represent a more acute or compensatory process compared to the food-location memory (CA3) and pattern separation (CA4) functions that showed durable effects. Notably, CA1 was the only subfield to show a non-significant but marginal association in the IS group at 24 months. This unexpected finding may reflect the distinct functional role of CA1 in reward valuation. CA1 conveys stronger value-related signals than CA3 and is specifically implicated in incremental value learning during foraging,^17,18^ a process that may operate through different mechanisms than the food-location memory and loss-of-control eating processes that characterize CA2/3 and CA4. The divergent patterns across subfields highlight that the hippocampal contribution to food-seeking behavior is not uniform and reflects the distinct computational roles of individual subfields.

The vulnerability of CA2/3 and CA4 to insulin resistance-related effects may additionally reflect the prolonged developmental time course of these subfields. Cross-sectional data suggest that CA2/3 and CA4 volumes increase steeply in early childhood and continue to increase through age 13-15 before stabilizing.^33^ However longitudinal data shows that CA2/3 and CA4 volumes showed linear volume decreases from childhood to adulthood, while CA1 showed nonlinear trajectories with early increases.^34^ This discrepancy may reflect methodological differences between these cross-sectional and longitudinal designs and subfield segmentation differences (e.g., using different FreeSurfer versions). Despite these differences, both studies converge on the conclusion that CA2/3 and CA4 undergo substantial structural change during adolescence. Tamnes et al.^34^ additionally found that general cognitive ability was positively associated with CA2/3 and CA4 volumes, reinforcing the key relation between neural volume and subsequent functional health of these subfields during development. This extended period of structural CA2/3 and CA4 maturation during adolescence may then be particularly sensitive to metabolic dysfunction. If insulin resistance disrupts the neurogenesis and synaptic plasticity processes that drive hippocampal volumetric change during adolescence, the resulting structural deficits may generate long-term functional consequences for food-reward processing and food-related contextual memory during a critical developmental period which may further worsen during typical hippocampal volume decline in adulthood.

While prior work helps us theorize why smaller hippocampal volume may predict greater unhealthy food-seeking over time in insulin-resistant youth, it is unclear why *larger* hippocampal volumes would linearly predict *reduced* food-seeking behaviors in youth also with untreated insulin resistance. One possible explanation could be that insulin-resistant youth who show improved food-seeking trajectories may deploy compensatory changes in hippocampal volume and plasticity to counteract the impact of insulin resistance on the hippocampus. In other words, rather than insulin resistance uniformly driving hippocampal volume reduction, some may exhibit greater neuronal resilience that preserves hippocampal volume and supports improved cognitive health outcomes. Supporting this theory, it has been demonstrated that hippocampal-specific insulin resistance can impair neuroplasticity independent of peripheral insulin resistance, and restoration of brain insulin activity can ameliorate cognitive deficits.^7^ It remains unclear, however, whether such compensatory changes would reflect a form of durable and long-term resilience or whether such compensation would instead reflect a form of short-term and unsustainable neural compensation that may lead to long-term neural exhaustion, subsequent atrophy and declined functioning.

These results identify a vulnerable population of youth who are obese, depressed, and insulin resistant but not yet diabetic, in whom metabolic dysregulation is likely to go undetected and untreated. Smaller hippocampal subfield volumes in insulin-resistant youth predicted increases in unhealthy food-seeking behavior over 24 months, a pattern that was not observed in insulin-sensitive youth. This suggests that the neurobehavioral consequences of insulin resistance may precede and potentially accelerate the trajectory toward clinical diabetes and worsening comorbidities such as obesity. Furthermore, the identification of insulin resistance as a moderator of hippocampal-behavioral trajectories points to insulin sensitivity as a potential intervention target with downstream behavioral implications. GLP-1 receptor agonists (GLP-1RAs) such as semaglutide and liraglutide are now FDA approved for chronic weight management in pediatric patients aged 12 and older with obesity (https://www.fda.gov/drugs/news-events-human-drugs/fda-approves-weight-management-drug-patients-aged-12-and-older). In addition to peripheral metabolic effects, GLP-1RAs act centrally through reward and appetite circuits; GLP-1 receptors are expressed across the hippocampus, ventral tegmental area, and nucleus accumbens.^16^ In the hippocampus specifically, GLP-1 receptor signaling has been shown to enhance learning and memory, promote neurogenesis, and decrease neuroinflammation.^35^ During et al.^35^ also demonstrated that GLP-1R- deficient mice have learning deficits that are restored after hippocampal GLP-1R gene transfer, and that overexpressing GLP-1R in the hippocampus improves learning and memory, providing direct evidence for a hippocampal-specific GLP-1R mechanism. Given that the hippocampal subfields identified in our study (CA2/3, and CA4) are regions where GLP-1 receptors are expressed and where insulin signaling is critical for neuronal metabolism and plasticity, GLP-1RAs represent a biologically plausible pharmacological strategy that could address both metabolic dysregulation and its neurobehavioral consequences. No pediatric trials have examined the effects of GLP-1RAs on hippocampal structure, food-seeking behavior, or these insulin resistance-brain-behavior mechanistic pathways. Whether GLP- 1RAs may improve insulin dynamics through interactive mechanisms, or whether GLP-1RAs may improve cognition through other mechanisms without addressing the negative impact of insulin resistance, however, remains unclear and requires future research.

Current American Academy of Pediatrics (AAP) and American Diabetes Association (ADA) guidelines recommend screening for abnormal glucose metabolism in youth ages 10 or pubertal onset with obesity and additional risk factors.^5,6^ These screening frameworks, however, rely on glucose-based diagnostic thresholds (fasting plasma glucose, 2-hour OGTT glucose, or HbA1c) and do not characterize insulin resistance itself. Using the OGTT-derived Matsuda ISI, we identified a population of insulin-resistant but non-diabetic youth who already exhibit structural neural changes predictive of escalating unhealthy food-seeking behavior, which may exacerbate obesity and comorbidities over time. Critically, when regressing out variance from peripheral metabolic factors, such as HOMA-IR, Insulin Area Under the Curve, fasting insulin, and insulinogenic index, respectively, our findings remained significant. This is consistent with evidence that HbA1c has limited sensitivity for detecting glucose intolerance in adolescents and may be insufficient to diagnose early T2DM.^36,37^ In our study here, insulin sensitivity groups were defined using a median split approach which coincided with established criteria for insulin resistance. Although insulin sensitivity moderated the hippocampal-food-seeking relation on a continuous scale, further categorical grouping of insulin sensitivity revealed that the relation was driven by individual differences for those in the Insulin Resistant group, with no predictive relation found for those in the Insulin Sensitive group. This suggests that the defined Matsuda ISI threshold effectively stratifies youth into neurobiologically meaningful vulnerability categories, rather than reflecting a linear dose-response relation. This is consistent with the concept of a threshold effect, wherein the neurobehavioral consequences of insulin resistance emerge once a critical level of metabolic dysregulation is reached. The specific threshold used in this study aligns with prior work establishing clinically meaningful Matsuda ISI thresholds. Radikova et al.^38^ proposed a Matsuda ISI cut-off of 50.0 for identifying insulin-resistant participants in a general adult population, while Frigerio et al.^39^ identified a Matsuda ISI 3.33 as optimal for capturing metabolic syndrome in adults who are overweight or obese. Our Matsuda ISI median split of 3.71, and the average Matsuda ISI within the Insulin Resistant group of 2.43, align with such clinical criteria proposals. Notably the AAP clinical practice guidelines state that fasting insulin is not a reliable measure of insulin resistance,^5^ further supporting the use of OGTT-derived measures such as the Matsuda ISI for identifying neurobehaviorally meaningful insulin resistance.

It is unclear whether our moderation findings are a function of peripheral or central insulin resistance, or both. Hippocampal insulin resistance can occur independently of peripheral insulin resistance and glycemic control, with intranasal insulin administration decreasing hunger through hippocampal-prefrontal cortex connections.^22^ Hippocampal-specific insulin receptor knockdown in rats has been shown to impair spatial learning and synaptic plasticity without affecting body weight, adiposity, or peripheral glucose homeostasis, establishing that central insulin resistance can produce neurocognitive effects independent of peripheral metabolic status.^7^ Our findings are preserved even after regressing out several factors which account for different peripheral insulin and glucose dynamics, suggesting that our results could be primarily driven by central insulin dynamics.

The present sample is comprised of and limited in interpretation to youth with obesity and moderate-to-severe depression, raising the question of whether these findings generalize to non-obese or non-depressed populations. BMI was controlled for in all analyses, and depression controlled in subsequent exploratory analyses, suggesting that the observed effects cannot be fully explained as a function of adiposity or depression severity. Given all individuals were in the top 98^th^ percentile of BMI and with moderate-to-severe depression, however, our individual differences of BMI and depression severity are largely constrained such that variance explained by these factors may still be detectable in a cohort with greater individual variability.

In conclusion, we consolidate and highlight several lines of future research we find warranted: (1) clinical trials integrating neuroimaging, behavioral outcomes, and GLP-1RA treatment in insulin-resistant youth are needed to determine whether improving insulin sensitivity can alter hippocampal-behavioral trajectories. (2) functional connectivity analyses and task-based fMRI paradigms targeting food-reward processing would clarify whether insulin resistance differentially affects hippocampal subfield-specific circuits, particularly the CA1-to-mPFC inhibitory pathway versus CA2/3-mediated memory consolidation processes. (3) studies using intranasal insulin paradigms or central insulin biomarkers could directly test whether our findings reflect central rather than peripheral insulin resistance. (4) longitudinal studies with repeated neuroimaging across multiple timepoints from childhood to adulthood would help determine whether the hypothesized compensatory hippocampal mechanisms in insulin-resistant youth (i.e., larger hippocampal volume at baseline predicted improvements in unhealthy food-seeking over time) are long-term and durable or are instead short-term and transient. (5) replication in a broader cohort with greater individual differences in BMI and depression measures is needed to determine whether insulin-resistant youth in general show this hippocampal-food-seeking trajectory relation, or if this relation instead is specific to depressed and obese youth. (6) investigation of the unexpected CA1 association in the insulin-sensitive group at 24 months, including whether the distinct role of CA1 in reward valuation operates through mechanisms independent of metabolic status.

## Methods

### Study Participants and Screening Criteria

The Measuring Obesity and Mood Effects On Neurobehaviors Through Maturation (MOMENTUM) study was a longitudinal prospective cohort study examining overweight, unmedicated youth ages 9 to 17 who were seeking treatment for depression. From a larger cohort of youth recruited from pediatric mood and weight control programs and community advertisements, data from 80 participants were included in this analysis. All participants provided written assent, and at least one parent or guardian provided written informed consent prior to all study procedures. The study was approved by the Stanford University Institutional Review Board. Secondary analyses of this data were approved by the UC Davis Institutional Review Board (IRBNet # 2303422-1).

Participants were evaluated on levels of depression severity using the Children’s Depression Rating Scale-Revised (CDRS-R)^40^ by a study clinician or trained coordinator. Individuals were included if: (1) their body mass index was at the 85^th^ percentile or higher based on the CDC BMI calculator for children and teens; (2) depression severity was categorized as being at least moderate to severe defined by a CDRS-R score of 35 or greater; (3) they were treatment seeking for active unremitting symptoms of depression; (4) they were unmedicated for depression; (5) they were generally early in the course of their illness.

Participants were excluded from the study if: (1) they were already being treated for a mood disorder when evaluated at screening visit; (2) they had type 1 or type 2 diabetes; (3) were taking medication that affected their mood, weight, or metabolism at the time of screening; (4) they had a contraindication for MRI (e.g. metal in their body or anterior-posterior diameter > 46 cm); or (5) their Full-4 IQ score on the Wechsler Abbreviated Scale of Intelligence (WASI)^41^ was < 70. Following recruitment, participants were assessed for current and lifetime psychiatric disorders using the Kiddie Schedule for Affective Disorders and Schizophrenia-Present and Lifetime version (KSADS-PL).^42^

Of the 80 participants analyzed at baseline, nine participants were excluded from analyses for missing baseline hippocampal or total brain volume measures. An additional four participants were excluded from analyses for missing Matsuda ISI measures necessary for our moderation analysis. Of the 67 participants who completed behavioral and MRI measures, 52 returned for 6-month follow-up and 33 returned for 24-month follow-up. Finally, one participant was removed from analyses as an outlier whose hippocampal volume was greater than three standard deviations from the mean. Thus, 51 and 32 participants were retained for subsequent analyses at 6- and 24-months follow-up, respectively.

### Anthropometric Measurement

At screening visits, height (accuracy of 0.1 cm) and weight (accuracy of 0.1 kg) were measured after the removal of shoes and jacket with the Seca 284, an electronic measuring station. Height was measured twice and then averaged. Pubertal status was assessed using the self-report Pubertal Development Scale^43^ in conjunction with clinician’s physical examination of secondary sex characteristics to confirm the self-reported Tanner stage ratings.

### Metabolic Measurement

Youth also completed a 2-hour oral glucose tolerance test (OGTT) to determine their insulin resistance. After 10 hours of fasting and an initial fasting blood draw, participants were given 75g bolus of oral glucose and had their blood drawn in 30-minute increments for 2 hours to measure insulin and glucose levels. Insulin values were determined using immunoassay.

Insulin sensitivity was assessed using the Matsuda Insulin Sensitivity Index (ISI), derived from the 2-hour glucose tolerance test (OGTT). The Matsuda ISI correlates strongly with the hyperinsulinemic-euglycemic clamp, the gold standard for measuring insulin sensitivity, and is superior to fasting-only indices such as HOMA-IR for detecting early metabolic dysregulation.^44^ The Matsuda ISI was calculated from results from the OGTT using the following formula: Matsuda ISI=10,000/(G_0_xI_0_xG_mean_xI_mean_)^1/2^. Higher Matsuda ISI values indicate less whole-body resistance to insulin (i.e., more insulin sensitive) while lower values indicate greater whole-body insulin resistance.

Participants were classified as insulin resistant or insulin sensitive using a median split of the baseline Matsuda ISI sample distribution for participants retained for our 6-month follow-up analyses (n=51), which aligned with established Matsuda ISI thresholds for insulin resistance of 50.0^38^ and 3.33.^39^ As described in the results, our median split of 3.71 and average Matsuda ISI of 2.43 in the Insulin Resistant group and 5.43 in the Insulin Sensitive group confirms that our insulin sensitivity groups align with proposed clinical guidelines.

### Food-Seeking Behavior

To quantify the motivation to obtain unhealthy food, we used a computerized adaptation of the Relative Reinforcing Value (RRV) task developed by Epstein and colleagues.^25^ The RRV paradigm operationalizes food reinforcement as the amount of work an individual is willing to perform to obtain food under a progressive-ratio schedule, in which response requirements increase across successive trials. This approach is grounded in behavioral choice theory and has been validated across developmental stages, with higher food reinforcement consistently associated with greater energy intake and higher BMI in children and adults.^25^ Prior to the task, participants selected one healthy snack and one unhealthy snack from an array of options. During the task, participants completed a computerized slot-machine like game in which they clicked a mouse to earn points redeemable for either their chosen healthy or unhealthy snack, with the healthy snack serving as the concurrent alternative reinforcer. Response requirements increased progressively across sequential trials, such that each successive food reward required more effort to obtain. Participants could allocate their effort toward either snack type and continued until they no longer wished to play. The total number of points earned for the unhealthy snack served as the primary outcome measure, with a greater number of points reflecting stronger motivation to obtain unhealthy food (i.e., greater unhealthy food-seeking behavior).

### Neuroimaging Data Acquisition

MRI data were originally collected and pre-processed in the prior laboratory of Dr. Manpreet Singh at Stanford University to which we report the methods of below. Neuroimaging data were acquired from all participants at the baseline visit. After participants were familiarized with the scanning environment in an MRI simulator, whole-brain images were acquired on a 3T GE Signa Excite scanner (General Electric Co., Milwaukee, WI) equipped with an 8-channel head coil. High-resolution structural images were collected using a fast spoiled gradient recalled (3D FSPGR) pulse sequence with the following parameters: TR=8.5 ms, TE=3.32 ms, TI=400 ms, flip angle=15°, field of view (x)=25.6 cm, matrix of 256 x 256, number of slices=186 slices in the axial plane, and a slice thickness of 1 mm.

Segmentation of the hippocampus was obtained with the publicly available FreeSurfer image analysis suite, version 5.3.^45^ Total hippocampal volume (mm^3^) was computed by summing the left and right hippocampus volumes. The automated recon-all pipeline, which includes intensity normalization, skull stripping, and segmentation of gray/white matter and subcortical structures, was completed for each scan. To ensure accuracy of grey/white matter segmentation and the inclusion of brain tissue, a trained investigator (MP) performed quality checks, while blinded to the clinical status of each participant. Manual corrections were performed by this rater where appropriate in accordance with previously established procedures.^46^

### Statistical analyses

All statistical analyses were conducted in R (version 4.3.2). Hippocampal and unhealthy food-seeking measures exceeding 3 standard deviations from the mean were excluded from analyses. As mentioned above, one participant yielded hippocampal volumes that exceeded three standard deviations from the mean and was excluded from subsequent analyses. Thus, we retained 51 participants for 6- month analyses and 32 participants for 24-month analyses. All models included sex, age, BMI percentile, total brain volume, and pubertal development score as covariates. Benjamini-Hochberg false discovery rate (FDR) correction was applied to control for multiple comparisons across hippocampal subregions.

### Correlation Comparisons

#### No Moderation

We first examined whether there exists a relation between hippocampal volume at baseline and change in unhealthy food-seeking (6mo – baseline; 24mo – baseline, respective) without insulin sensitivity as a moderator. Linear regression modeling using Ordinary Least Squares (OLS) was applied using the following model parameters shown in equation (1):

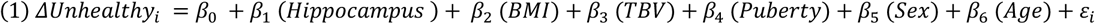

Our ROI predictor (*β*_1_) in the following model applied average hippocampal volume as well as hippocampal subregions CA1, CA2/3, and CA4, respectively. We additionally applied Bayesian Inference as additional evidence in support of or evidence in contrast to our frequentist results. We computed the unadjusted Bayes Factor (BF_10_) using the BayesFactor() package in R to examine the degree of evidence in favor of the alternative hypothesis (i.e., a true correlation) versus the null hypothesis (i.e., no meaningful relation between factors) using the following scale:

#### With Continuous Insulin Sensitivity Moderation

We examined whether insulin sensitivity moderated the relation between hippocampal volume at baseline and change in unhealthy food-seeking (6mo – baseline; 24mo – baseline, respective) by applying insulin sensitivity as a continuous moderator. Insulin sensitivity was added as a continuous moderator by using individual differences in Matsuda ISI. To reduce multicollinearity, insulin sensitivity and ROI predictors were mean-centered prior to analyses.

Linear regression modeling using Ordinary Least Squares (OLS) was applied using the following model parameters shown in equation (2):

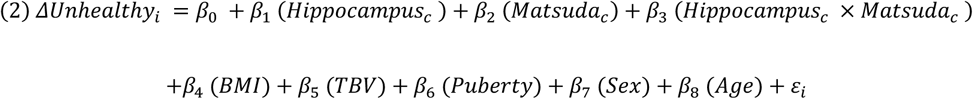

Like the previous model, our ROI predictor (*β*_1_) in the following model applied average hippocampal volume as well as hippocampal subregions CA1, CA2/3, and CA4, respectively. We applied Bayesian Inference to directly evaluate evidence in favor of an interaction as additional evidence to our frequentist results. We computed a covariate-adjusted Bayes Factor (BF_10_) comparing the full model to a reduced model retaining only the main effects and covariates (i.e., no interaction term) using the lmBF() function in R.

#### With Categorical Insulin Sensitivity Moderation

We next examined whether categorical insulin sensitivity criteria aligned with clinical patients with insulin resistance would demonstrate a neurophysiologically-meaningful phenotype for predicting unhealthy food-seeking trajectories in the hippocampus. This model, i.e., equation (3), has a similar structure as the continuous moderation model described above, with the following modifications. Participants were classified as either insulin resistant or insulin sensitive using a median split method on Matsuda ISI of 3.71. As described above, this median split value is aligned with proposed clinical criteria using Matsuda ISI to identify insulin resistance. The model was first fit to the Insulin Sensitive group as the reference group followed by refitting to the Insulin Resistant group as the reference group.

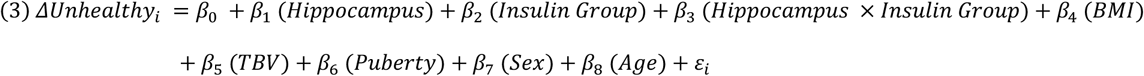

Given it is unclear how covariate-adjusted BF_10_ performs when the number of model parameters approaches the sample size (i.e., 6-month: IR n=25, IS n=26; 24-month: n=16 per group),^47^ we calculated a simple, unadjusted BF_10_ for each insulin sensitivity group (IR, IS) to examine additional evidence for the core relation between hippocampal volume and unhealthy food-seeking trajectories.

#### Financial Disclosures for Manpreet K. Singh, MD MS

Dr. Singh has received research support from the National Institutes of Health, the Patient Centered Outcomes Research Institute, Advanced Neuromodulation Systems/Abbott, AbbVie Inc, Alto Neuroscience, and IntraCellular Therapies/Johnson & Johnson. She has been on a data safety monitoring board for a study funded by the National Institute of Mental Health. She has in the past 3 years consulted for or been on an advisory board for AbbVie Inc., Alkermes, Almatica, Alto Neuroscience, Axsome, Boehringer-Ingelheim, Bristol Myers Squibb, Johnson and Johnson, Karuna Therapeutics, Neumora, and Supernus. She receives honoraria from the American Academy of Child and Adolescent Psychiatry and royalties from American Psychiatric Association Publishing and Thrive Global.

The authors declare no other competing interests or disclosures.

## Data Availability

All data produced in the present study may be requested with reasonable request to the authors and corresponding UC Davis Health approvals and security clearances.

## Notes

### Competing Interest Statement

The authors have declared no competing interest.

### Author Declarations

Institutional Review Board of University of California, Davis gave ethical approval for this work (IRBNet # 2303422-1).

### Funding

- National Institute of Mental Health (NIMH): R01 MH106581, R01 MH106581S1

