## Supplemental Materials for "Hippocampal Volume Predicts Unhealthy Food-Seeking Trajectories in Insulin-Resistant, but Not Insulin-Sensitive, Youth with Obesity and Depression"

#### 1. Depression Analyses - 6 Months

We first determined whether change in depression severity, a comorbidity of obesity, predicts change in unhealthy food-seeking from baseline to 6 months, or whether these two measures are dissociable. We found change in depression severity did not predict change in unhealthy food-seeking from baseline to 6 months ($p = .69$).

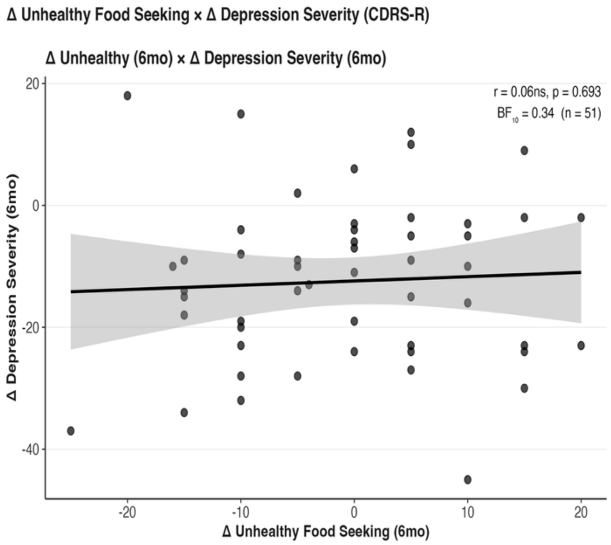


***Figure 1.*** Pearson correlation comparison of change in unhealthy food seeking from 6 months – baseline (x-axis) and change in depression severity from 6 months – baseline (y-axis). Top right: correlation coefficient (r), p-value, $BF_{10}$, and sample size (n).

#### Continuous Insulin Sensitivity Moderation

We next included depression severity as an additional covariate to our existing continuous insulin sensitivity model. Individual differences of insulin sensitivity significantly moderated the relation between hippocampal volume at baseline and change in unhealthy food-seeking behavior at 6-month follow-up ($p < .05$), such that as insulin sensitivity decreases the relation between hippocampal volume and change in unhealthy food-seeking becomes more negative. This moderation was significant across cornu ammonis hippocampal subfields CA1, CA2/3, and CA4 at 6- and 24-month follow-up (all $p < .05$) and remained predictive following Benjamini-Hochberg FDR correction for multiple comparisons at 6- and 24-month follow-up (all $p_{BH} < .05$).

### Categorical Insulin Sensitivity Moderation

We next included depression severity as an additional covariate to our existing insulin group model. Average hippocampal volume at baseline did not significantly predict changes in unhealthy food-seeking from baseline to 6-months among the IR group (but marginal; p=.06) or IS group (p=.91). The interaction between insulin sensitivity group and hippocampal volume was significant (p < .05).

Within the cornu ammonis hippocampal subfield, baseline volumes for subfields CA2/3, and CA4 were significantly predictive of change in unhealthy food-seeking behavior only among IR participants (all p < .05) and remained predictive following Benjamini-Hochberg FDR correction for multiple comparisons (all pBH < .05) with significant interaction (all pBH < .05), such that smaller hippocampal subfield volumes at baseline predicted increases in unhealthy food-seeking for IR but not IS groups.

### Predictors of Change in Depression Severity

We next examined the role of insulin sensitivity on the relation between hippocampal volume and change in depression severity from baseline to 6 months to further verify whether comorbid depression could explain our primary findings with change in unhealthy food-seeking. We found no significant relation between hippocampal volume (and subregions, respectively) at

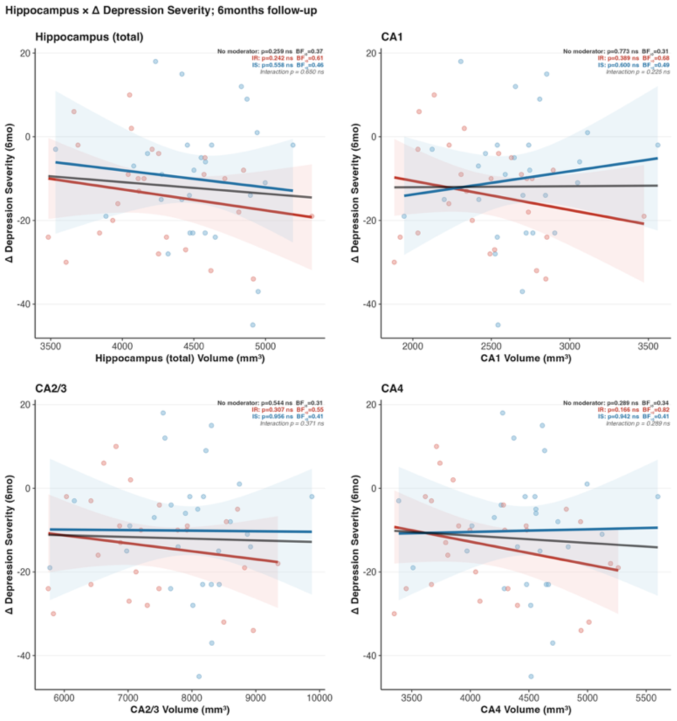


***Figure 2.*** Moderation of Insulin Sensitivity on the relation between hippocampal volume at baseline and change in depression severity from 6-months – baseline. Plots reflect simple slopes with displayed values reflecting addition of covariates. Red line and text: Insulin Resistant group; Blue line and text: Insulin Sensitive group. For each comparison, the main effect p, Bayes Factor, and interaction p are presented in the top right side of the comparison plot. We present four comparisons using the following regions of interest (x-axis): average hippocampal volume (top left), and hippocampal subfields CA1 (top right), CA2/3 (bottom left), and CA4 (bottom right). Dashed line: simple slope without insulin sensitivity moderation (all p > .05).

baseline and change in depression severity from baseline to 6 months for youth in the Insulin Resistant or IS group (all $p < .05$).

2. **Depression Longitudinal Validation – 24 Months**

**Depression Severity x Unhealthy Food Seeking Trajectories**

We first determined whether change in depression severity predicts change in unhealthy food-seeking from baseline to 24 months, or whether these two measures are dissociable. Please note that one participant did not have a depression severity score at 24-month follow-up, thus the 24-month sample size was reduced by one from 32 to 31. We found change in depression severity did not predict change in unhealthy food-seeking from baseline to 24 months ($p = .61$).

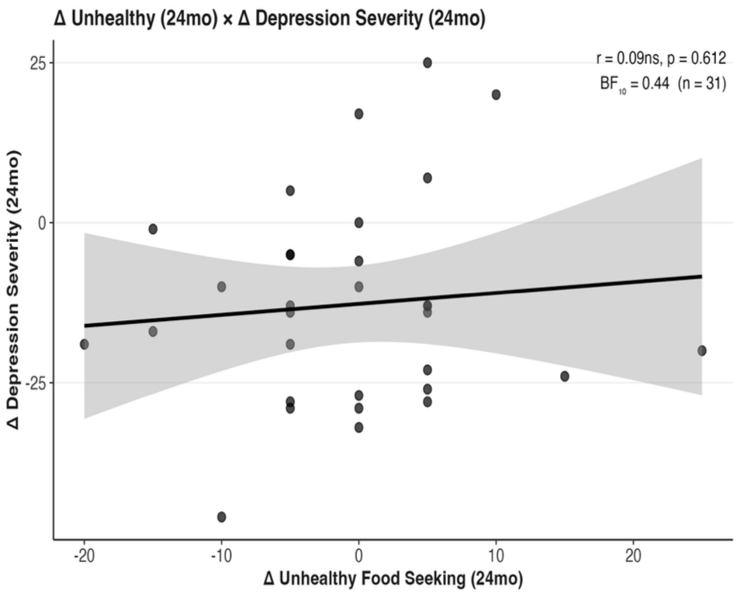


***Figure 3***. Pearson correlation comparison of change in unhealthy food seeking from 24 months – baseline (x-axis) and change in depression severity from 24 months – baseline (y-axis). Top right: R, p-value, $BF_{10}$, and sample size key.

**Continuous Insulin Sensitivity Moderation**

Individual differences of insulin sensitivity significantly moderated the relation between hippocampal volume at baseline and change in unhealthy food-seeking behavior at 24-month follow-up ($p < .05$), such that as insulin sensitivity decreases the relation between hippocampal volume and change in unhealthy food-seeking becomes more negative. This moderation was significant across cornu ammonis hippocampal subfields CA1, CA2/3, and CA4 at 6- and 24-month follow-up (all $p < .05$) and remained predictive following Benjamini-Hochberg FDR correction for multiple comparisons at 6- and 24-month follow-up (all $p_{BH} < .05$).

**Categorical Insulin Sensitivity Moderation**

Using categorical grouping of insulin sensitivity via median split, average hippocampal volume at baseline did not significantly predict change in unhealthy food-seeking baseline to 24-months follow-up neither for participants in the IR group ($p = .44$) nor for participants in the IS group ($p = .42$). The interaction was additionally non-significant ($p = .08$). We further found that baseline volumes for subregions CA1, 2/3, and 4 were non-significant, but CA2/3 marginally predictive of 6-month change in unhealthy food-seeking only for participants in the IR group (CA1: $p = .45$; CA2/3: $p = .056$; CA4: $p = .08$). CA1 remained significant ($p < .05$) whereas CA2/3 and CA4 was non-significant for participants in the IS group ($p > .05$). CA1 remained predictive following Benjamini-Hochberg FDR correction for multiple comparisons ($p_{BH} < .05$). Although the overall significance was mostly lost, subfield relations retained or trended towards our primary findings. What remains unclear is whether this reduction in relation significance reflects a significant proportion of variance accounted for by baseline depression severity that is clinically meaningful given our modest sample size at 24-month follow-up, which limits such interpretation after adding an additional exploratory covariate. Further investigation is needed to better understand the role of depression severity as a covariate in this model.

**Predictors of Change in Depression Severity**

We again examined the relation between hippocampal volume and change in depression severity across insulin sensitivity groups to further verify whether comorbid depression could explain our primary findings with change in unhealthy food-seeking from baseline to 24 months. We found no significant relation between hippocampal volume (and subregions, respectively) at baseline and change in depression severity from baseline to 24 months for youth in the Insulin Resistant or IS group (all $p < .05$).

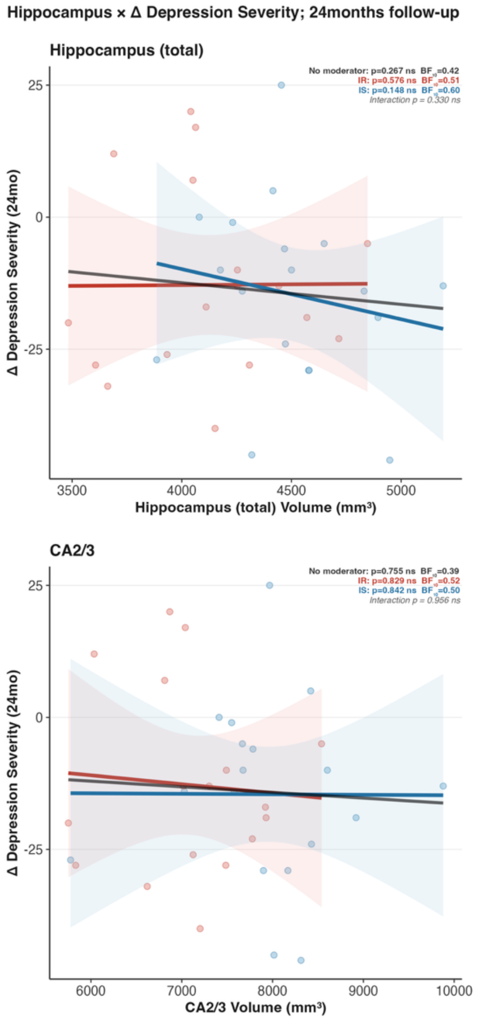


***Figure 4.*** Moderation of Insulin Sensitivity on the relation between hippocampal volume at baseline and change in depression severity from 6-months – baseline. Plots reflect simple slopes with displayed values reflecting addition of covariates. Red line and text: Insulin Resistant group; Blue line and text: Insulin Sensitive group. For each comparison, the main effect p, Bayes Factor, and interaction p are presented in the top right side of the comparison plot. We present four comparisons using the following regions of interest (x-axis): average hippocampal volume (top left), and hippocampal subfields CA1 (top right), CA2/3 (bottom left), and CA4 (bottom right). Dashed line: simple slope without insulin sensitivity moderation (all $p > .05$).
